# Beyond Genotype: Multidomain Biomarker-Integrated Risk Scores Reveal Prognostic phenotypes Among Individuals with High-Risk *APOL1* Genotypes

**DOI:** 10.64898/2026.06.03.26354877

**Authors:** George Vasquez-Rios, Kinsuk Chauhan, Nidhi Naik, Pattharawin Pattharanitima, Lili Chan, Kirk N Campbell, Girish N. Nadkarni, Steven G. Coca

## Abstract

*APOL1* high-risk variants markedly increase susceptibility to kidney disease among individuals of African ancestry; however, only a subset of carriers develops clinically significant CKD or ESKD. This discrepancy highlights a gap between genetic risk and clinical trajectory. Current prognostic tools rely primarily on eGFR and albuminuria, which incompletely reflect the underlying biological processes driving *APOL1*-associated kidney injury. We hypothesized that plasma biomarkers reflecting inflammatory and tubular injury pathways could identify biologically active disease states within this genetically “high-risk” population and improve prognostic stratification.

**Methods:** Participants from the Mount Sinai Bio*Me* Biobank carrying two *APOL1* high-risk alleles (G1, G1; G1, G2; or G2 G2) were followed for a median of 6 years. Baseline plasma biomarkers of inflammation and tubular injury (TNFR1, TNFR2, KIM-1, MCP-1, YKL-40, IL-18, suPAR) were measured. The composite outcome was sustained ≥40% decline in eGFR or ESKD. Multivariable Cox models assessed associations between biomarkers and outcomes. A weighted biomarker risk score was derived from tertile-based hazard ratios and categorized into low-, moderate-, and high-risk groups.

**Results:** Among 498 participants (median eGFR 83 ml/min/1.73 m^2^), 80 (16.1%) reached the composite outcome. Higher concentrations of TNFR1, TNFR2, suPAR, KIM-1, and IL-18 were independently associated with kidney events after multivariable adjustment. Event rates were 7% in the low-risk group, 16% in the moderate-risk group, and 36% in the high-risk group.

**Conclusions:** Plasma biomarkers reflecting inflammatory and tubular injury pathways reveal marked heterogeneity in kidney outcomes among individuals with high-risk *APOL1* genotypes. Integration of these signals into a biology-weighted score identifies distinct prognostic phenotypes beyond genotype and traditional clinical measures, supporting multidomain biomarker frameworks for risk stratification and potential trial enrichment in *APOL1*-associated kidney disease.

## 1. Introduction

The discovery of apolipoprotein L1 (*APOL1*) risk variants fundamentally changed our understanding of excess kidney failure among individuals of African ancestry (1,2). However, an enduring paradox remains: although *APOL1* high-risk genotypes confer substantial susceptibility to kidney disease, only a minority of carriers develop chronic kidney disease (CKD) or end-stage kidney disease (ESKD) during their lifetime (1–3). This discrepancy underscores a critical limitation of genotype-based risk attribution—genetic risk alone does not define clinical trajectory.

Current clinical prognostication in CKD relies primarily on estimated glomerular filtration rate (eGFR) and albuminuria. While useful, these measures reflect late manifestations of kidney damage and do not capture the underlying biological processes driving disease progression (4). This limitation is particularly relevant in *APOL1*-associated kidney disease, where pathophysiology is shaped by complex interactions between podocyte stress, tubular injury, immune activation, and systemic inflammatory triggers (2,5,6). In addition, *APOL1*-associated kidney risk appears to be context dependent, with environmental and “second-hit” interactions increasingly recognized as key modifiers of disease expression (7–9).

Emerging experimental and translational data suggest that *APOL1* cytotoxicity leads to podocyte injury with secondary tubulointerstitial consequences, mediated in part by inflammatory and immune pathways (2,5,6). Yet, tools to clinically measure these biological domains and translate them into prognostic insight are lacking. Identifying individuals with biologically active disease states—rather than relying solely on genotype or static clinical markers—may be essential to improving risk stratification in this population.

In other areas of medicine, multidomain scoring systems that integrate biomarkers with clinical variables have transformed prognostication. For example, biomarker-enhanced cardiovascular risk models and composite molecular scoring systems in hematology have demonstrated how integrating biological signals into structured scoring systems can reveal clinically meaningful heterogeneity within populations otherwise considered uniformly at risk (10,11).

Biomarkers of tubular injury, inflammation, and repair—including TNFR1, TNFR2, KIM-1, MCP-1, YKL-40, IL-18, and suPAR—have demonstrated prognostic value across diverse kidney populations (12–20). Whether these biomarkers can identify biologically distinct prognostic phenotypes among individuals already labeled as “high-risk” due to *APOL1* genotype remains unknown (21). We therefore sought to determine whether a multidomain biomarker panel could identify evidence of biologically active disease within individuals carrying high-risk *APOL1* genotypes and whether integration of these signals into a composite risk score could stratify this population into clinically meaningful prognostic groups.

## 2. Methods

### 2.1 Study participants

This retrospective cohort study utilized DNA, plasma, and clinical data from participants enrolled in the Bio*Me* Biobank Program of the Charles Bronfman Institute for Personalized Medicine at the Icahn School of Medicine at Mount Sinai between 2007 and 2017. Bio*Me* is an Institutional Review Board (IRB)–approved, comprehensive biorepository linked to electronic medical records (EMR) and is notable for its high degree of ethnic diversity.^1^ □The biobank is fully integrated into clinical workflows and recruits participants directly from more than 30 clinical sites, including waiting areas and phlebotomy stations, through dedicated research staff. For the present analysis, we included only Bio*Me* participants with high-risk *APOL1* genotypes (G1/G1, G2/G2, or G1/G2). The Bio*Me* Biobank is an Institutional Review Board (IRB)-approved, a consented electronic medical record (EMR)-linked medical care setting biorepository in an ancestrally diverse local community of upper Manhattan.

### 2.2 Clinical data and baseline information

Demographic information, including age, sex, and self-reported African ancestry, was obtained from enrollment questionnaires. Clinical variables—including serum creatinine, hemoglobin A1c (HbA1c), urine protein-to-creatinine ratio (UPCR), and urine albumin-to-creatinine ratio (UACR)—were extracted from the EMR.

Baseline was defined as the clinical value closest to the Bio*Me* enrollment date within the preceding 12 months. Estimated glomerular filtration rate (eGFR) was calculated using the CKD-EPI creatinine equation. Median laboratory values were calculated within 3-month intervals and used for outcome ascertainment.

Body mass index (BMI) was calculated as weight in kilograms divided by height in meters squared (kg/m^2^). Hypertension and type 2 diabetes mellitus at baseline were defined using validated Electronic Medical Records and Genomics (eMERGE) phenotyping algorithms. Coronary artery disease and heart failure were identified using validated algorithms and ICD-9/10 codes. Participants were considered to be receiving angiotensin-converting enzyme inhibitors (ACEIs) or angiotensin II receptor blockers (ARBs) if an active prescription was documented at the time of enrollment.

Missing UACR values were imputed using the method described by Nelson et al.^1^ □for participants without diabetes and were assigned a value of 10 mg/g for those with diabetes. This conservative approach is consistent with CKD risk prediction strategies in which low-grade albuminuria is assumed when measurements are unavailable. UACR was included only as a covariate for multivariable adjustment and was not used in the definition of study outcomes. Follow-up time was calculated from the Bio*Me* enrollment date to the most recent clinical encounter documented in the electronic medical record.

### 2.3 *APOL1* genotyping

High-risk African American participants were identified through direct genotyping to determine ancestral (G0), G1, and G2 allele status of *APOL1*. Genotyping accuracy was validated through intra- and inter-assay precision testing using 48 positive and 10 negative control samples.^1^□ Genotypes were confirmed using Sanger sequencing, with complete concordance between sequencing and direct genotyping results. High-risk genotypes were defined as G1/G1, G2/G2, or G1/G2.

### 2.4 Biomarker measurements

Plasma samples were collected and aliquoted at the time of BioMe enrollment and stored at −80 °C until analysis. Baseline biomarker concentrations were measured from these stored samples.

Plasma soluble urokinase plasminogen activator receptor (suPAR) concentrations were measured using enzyme-linked immunosorbent assay (ELISA). Plasma IL-18, MCP-1, YKL-40, TNFR1, TNFR2, and KIM-1 concentrations were measured using multiplex cytokine arrays from Mesoscale Diagnostics. These methods have been described previously.

Investigators performing biomarker measurements were blinded to clinical outcomes. Intra-assay coefficients of variation (CVs) were 3.5%, 3.9%, and 4.5% for TNFR1, TNFR2, and KIM-1, respectively. Inter-assay CVs were 12.4%, 10.8%, and 7.7% for TNFR1, TNFR2, and KIM-1, respectively. The average lower limits of detection across assay runs were 12.5 pg/mL for TNFR1, 7.8 pg/mL for TNFR2, and 9.0 pg/mL for KIM-1. Given evaluation of multiple biomarkers and models, p values should be interpreted as exploratory and hypothesis-generating, with emphasis placed on effect sizes, consistency across adjusted models, and biological plausibility.

### 2.5 Outcomes and biomarker risk scores

The primary outcome was a composite kidney endpoint defined as the development of ESKD or a sustained decline in eGFR of ≥40% during follow-up (median 6 years). ESKD was defined as initiation of chronic dialysis or receipt of kidney transplantation and was ascertained through linkage with the United States Renal Data System (USRDS). A sustained ≥40% decline in eGFR was defined as a reduction of ≥40% from baseline confirmed at two or more measurements at least 3 months apart.

To translate the prognostic information contained within multiple biomarkers into a single interpretable measure, we constructed a biology-weighted biomarker risk score. For each biomarker, participants were first categorized into tertiles. Integer point values were then assigned to each tertile according to the magnitude of association observed in the fully adjusted Cox regression model (Model 3), such that biomarkers demonstrating stronger associations with kidney events contributed proportionally greater weight to the overall score (1 point for adjusted hazard ratios [aHR] 1.0 to <1.5; 2 points for aHR 1.5 to <2.0; 4 points for aHR 2.0 to <4.0; and 8 points for aHR ≥4.0).

For each participant, points across all biomarkers were summed to generate a composite weighted risk score reflecting the cumulative burden of abnormal biomarker signals across inflammatory and tubular injury domains. The distribution of total scores was divided into five ordered categories and subsequently consolidated into low-, moderate-, and high-risk strata for outcome analyses (**Figure 1**). Biomarkers were categorized into tertiles to create clinically interpretable groupings without assuming linear relationships, and integer weights based on adjusted hazard ratios were applied to preserve proportionality of risk while maintaining interpretability. Final score categories were defined according to the observed score distribution within the cohort to avoid arbitrary thresholds and reflect natural separation of risk.

**Figure 1.**
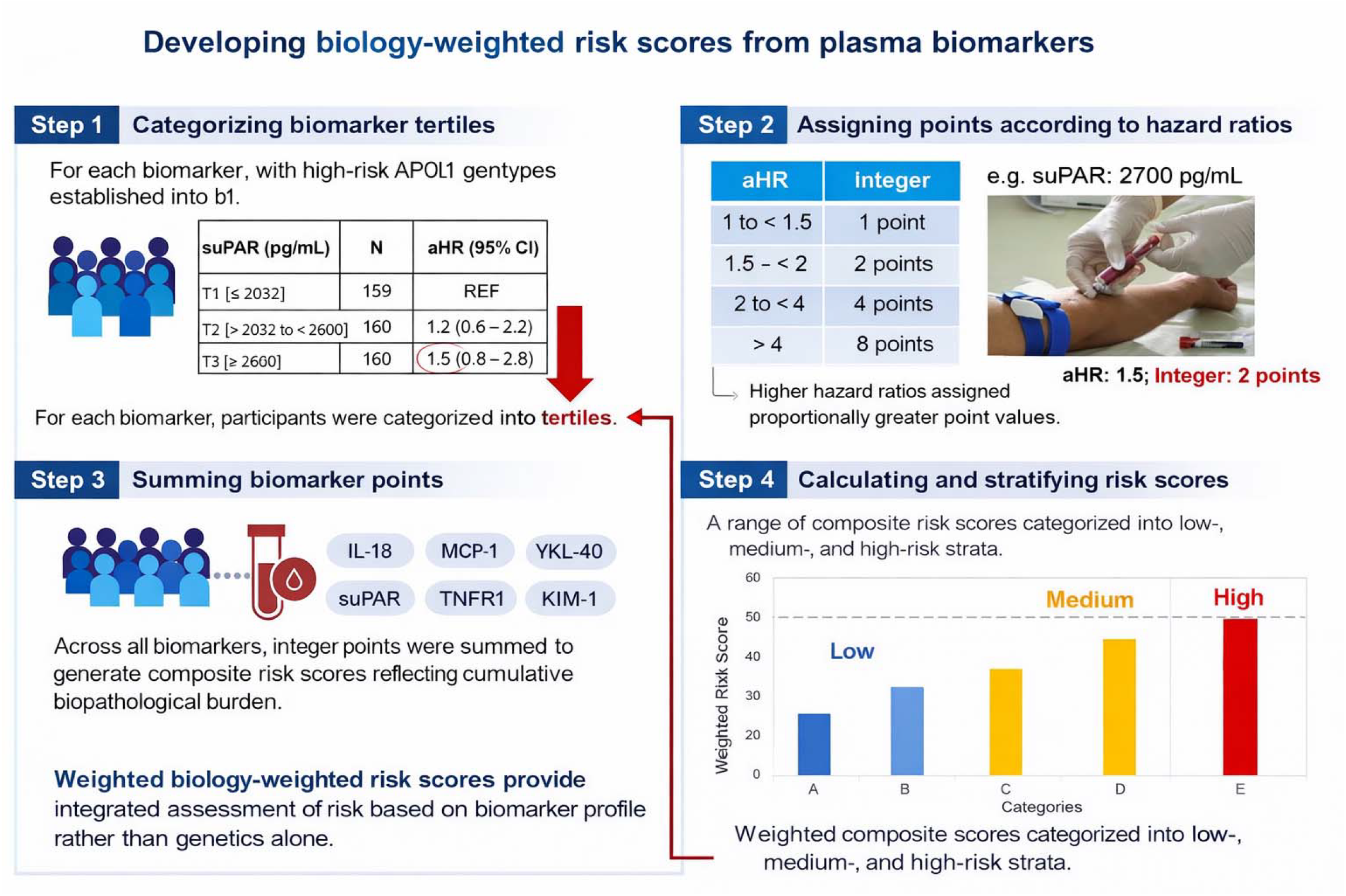
Development of a biology-weighted biomarker risk score for prognostic stratification in individuals with high-risk APOL1 genotypes. **Footnote:** This schematic illustrates the stepwise development of a composite, biology-weighted risk score derived from plasma biomarkers reflecting distinct pathophysiological domains of kidney injury. Step 1: Biomarker concentrations were divided into tertiles and corresponding adjusted hazard ratios (aHRs) for kidney events were calculated. Step 2: Integer points were assigned to each tertile based on the magnitude of the aHR. Step 3: Points across all biomarkers were summed to generate a composite risk score for each participant. Step 4: Composite scores were categorized into low-, moderate-, and high-risk strata according to their distribution. APOL1, apolipoprotein L1; aHR, adjusted hazard ratio; suPAR, soluble urokinase plasminogen activator receptor; TNFR1/2, tumor necrosis factor receptor 1 and 2; KIM-1, kidney injury molecule-1; MCP-1, monocyte chemoattractant protein-1; YKL-40, chitinase-3-like protein 1; IL-18, interleukin-18.

### 2.6 Statistical analysis

Baseline characteristics and biomarker concentrations were summarized using means with standard deviations or medians with interquartile ranges, as appropriate based on data distribution. Between-group comparisons were performed using Student’s t-tests or Wilcoxon rank-sum tests for continuous variables and chi-square tests for categorical variables. Associations between biomarkers and baseline clinical characteristics were assessed using Spearman partial correlation coefficients.

After verifying the proportional hazards assumption, Cox proportional hazards regression models were used to estimate hazard ratios (HRs) for the composite kidney outcome as a function of each biomarker. Biomarkers were analyzed both continuously after log_2_ transformation (interpretable as hazard per doubling of biomarker concentration) and categorically by tertiles. Three hierarchical models were constructed: Model 1 was unadjusted. Model 2 adjusted for age, sex, baseline eGFR, log_10_-transformed UACR, and body mass index (BMI). Model 3 further adjusted for type 2 diabetes mellitus, heart failure, systolic blood pressure, and use of angiotensin-converting enzyme inhibitors or angiotensin receptor blockers.

Time-to-event was defined as the time from BioMe enrollment to the first occurrence of dialysis, kidney transplantation, or sustained ≥40% decline in eGFR. Participants without events were censored at the end of the observation period. Discriminative performance of the models was assessed using Harrell’s concordance statistic (C-statistic) for time-to-event data. We calculated the C-statistic for a clinical model consisting of demographics, baseline eGFR, comorbidities, medication use, and log10-transformed UACR. We then recalculated the C-statistic after adding each biomarker individually and after adding all biomarkers simultaneously. Improvements in discrimination were evaluated by comparing changes in Harrell’s C-statistic across models. The C-statistic was internally validated using 1000 bootstrap resamples to account for optimism bias.

Model performance was assessed primarily in terms of discrimination. Calibration was not evaluated, as the objective was prognostic stratification and score-based enrichment rather than development of a clinical prediction model. All tests were two-sided, and p values <0.05 were considered statistically significant. Statistical analyses were performed using SAS version 9.4 (SAS Institute Inc., Cary, NC).

## 3. Results

### 3.1 Baseline characteristics of participants

Follow-up data and biospecimens were available for 498 Bio*Me* participants carrying two APOL1 risk alleles. The median age was 56 years, and 337 participants (68%) were women. The median baseline eGFR was 83 mL/min/1.73 m^2^ (IQR 69–99). During follow-up, 80 participants (16.1%) reached the composite kidney outcome.

Participants who experienced the outcome were older (66 vs. 55 years) and had lower baseline eGFR values (75.5 vs. 85.5 mL/min/1.73 m^2^) compared with those who did not reach the endpoint. They also had a higher prevalence of comorbid conditions, including type 2 diabetes mellitus, coronary artery disease, and heart failure. A detailed comparison of baseline characteristics between groups is presented in **Table 1**. Participants who reached the kidney outcome also had higher median concentrations of several biomarkers at baseline. Spearman partial correlations between biomarkers and key kidney parameters are shown in **Table 2**.

**Table 1:**
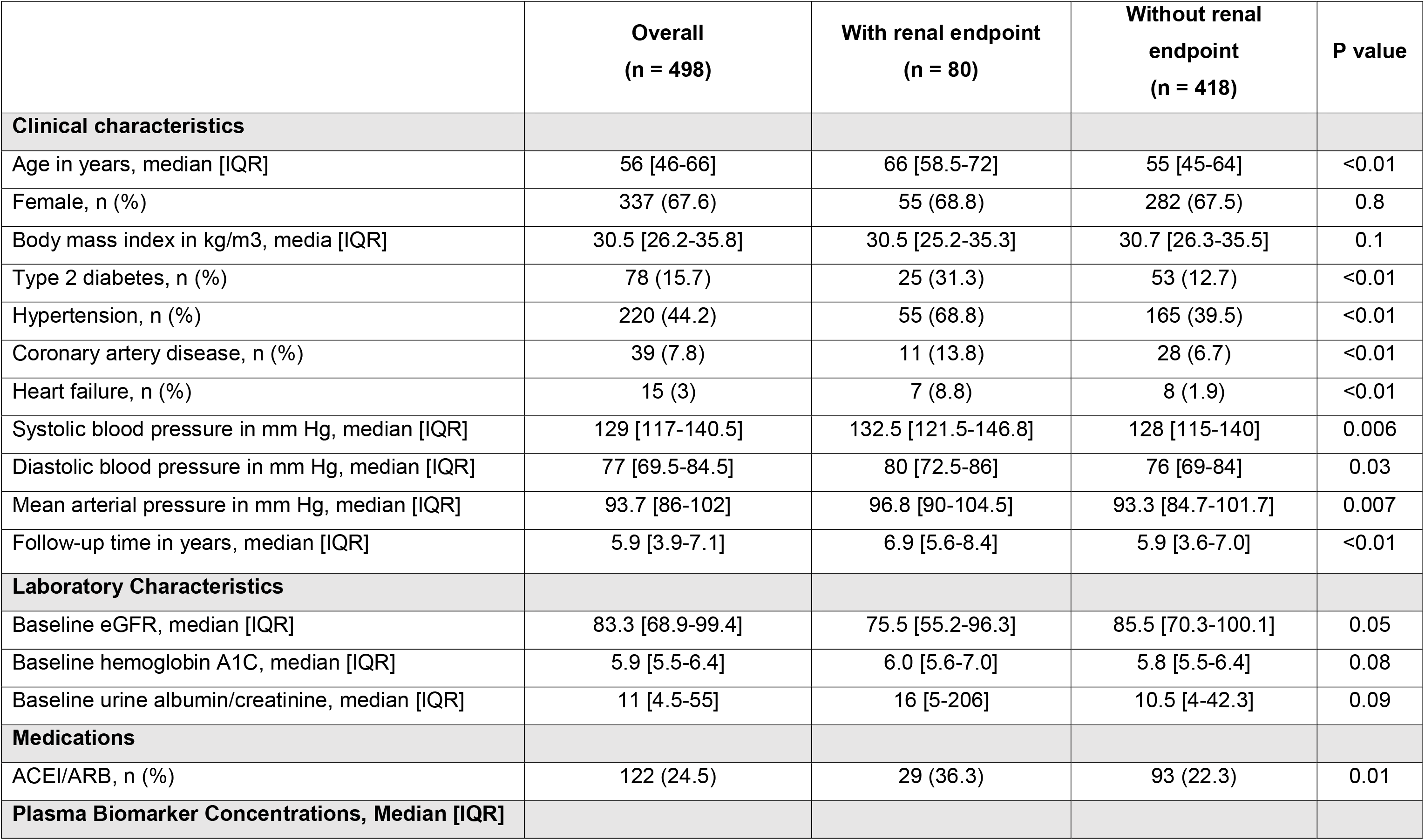
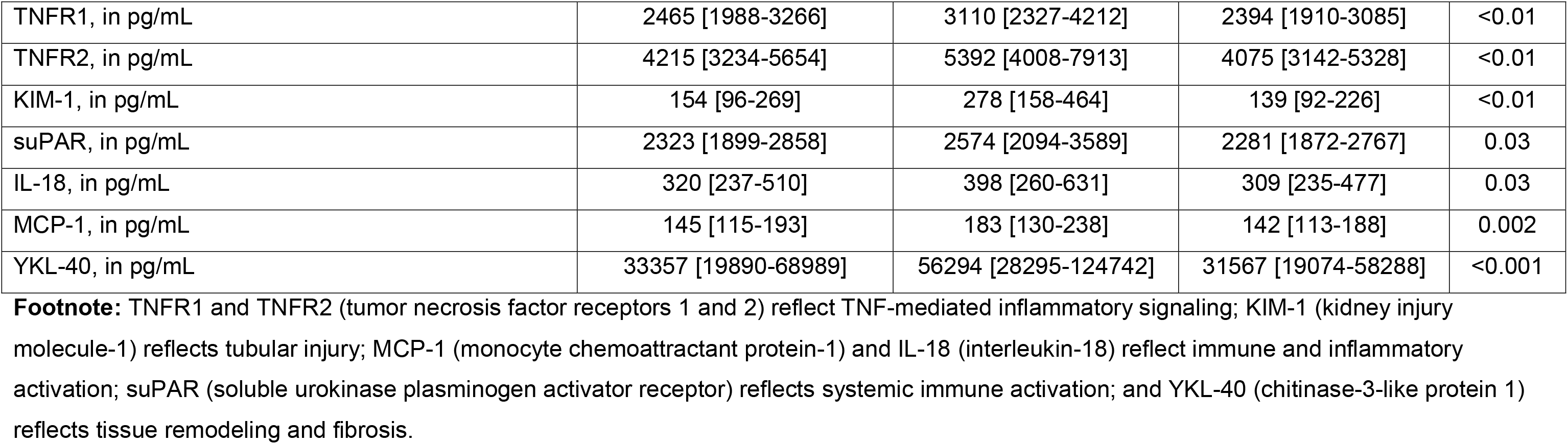
Baseline Characteristics of African American participants with the *APOL1* high-risk variants, with and without the kidney endpoint.

**Table 2:** Spearman correlations between plasma biomarkers and risk factors.

| Biomarker | suPAR | IL-18 | MCP-1 | YKL-40 | TNFR1 | TNFR2 | KIM1 | Age | eGFR | HbA1c | UACR-UPCR | BMI |
| --- | --- | --- | --- | --- | --- | --- | --- | --- | --- | --- | --- | --- |
| suPAR | NA | 0.3* | 0.3* | 0.4* | 0.5* | 0.6* | 0.4* | 0.3* | -0.3* | 0.1 | 0.1 | 0.2* |
| IL-18 | 0.3* | NA | 0.2* | 0.3* | 0.3* | 0.4* | 0.3* | 0.1* | -0.2* | 0.1* | 0.1 | 0.1* |
| MCP-1 | 0.3* | 0.2* | NA | 0.4* | 0.4* | 0.4* | 0.4* | 0.3* | -0.3* | 0.1* | 0.2* | 0.1 |
| YKL-40 | 0.4* | 0.3* | 0.4* | NA | 0.4* | 0.4* | 0.5* | 0.4* | -0.3* | 0.1 | 0.1 | 0.1* |
| TNFR1 | 0.5* | 0.3* | 0.4* | 0.4* | NA | 0.8* | 0.4* | 0.3* | -0.4* | 0.2* | 0.2* | 0.2* |
| TNFR2 | 0.6* | 0.4* | 0.4* | 0.4* | 0.8* | NA | 0.4* | 0.3* | -0.4* | 0.1* | 0.1 | 0.2* |
| KIM-1 | 0.4* | 0.3* | 0.4* | 0.5* | 0.4* | 0.4* | NA | 0.5* | -0.4* | 0.3* | 0.4* | 0.1 |
| Age | 0.3* | 0.1* | 0.3* | 0.4* | 0.3* | 0.3* | 0.5* | NA | -0.5* | 0.3* | 0.1 | -0.1 |
| eGFR | -0.3* | -0.2* | -0.3* | -0.3* | -0.4* | -0.4* | -0.4* | -0.5* | NA | -0.2* | -0.2* | -0.1 |
| HbA1c | 0.1 | 0.1* | 0.1* | 0.1 | 0.2* | 0.1* | 0.3* | 0.3* | -0.2* | NA | 0.2* | 0.1* |
| UACR | 0.1 | 0.1 | 0.2* | 0.1 | 0.2* | 0.1 | 0.4* | 0.1 | -0.2* | 0.2* | NA | -0.02 |
| BMI | 0.2* | 0.1* | 0.1 | 0.1* | 0.2* | 0.2* | 0.1 | -0.01 | -0.1 | 0.1* | -0.02 | NA |

\*Significant p-values
**Footnote:** suPAR, soluble urokinase plasminogen activator receptor; IL-18, interleukin-18; MCP-1, monocyte chemoattractant protein-1 (CCL2); YKL-40, chitinase-3-like protein 1; TNFR1, tumor necrosis factor receptor 1; TNFR2, tumor necrosis factor receptor 2; KIM-1, kidney injury molecule-1; eGFR, estimated glomerular filtration rate; HbA1c, hemoglobin A1c; UACR, urine albumin-to-creatinine ratio; UPCR, urine protein-to-creatinine ratio; BMI, body mass index. Biomarker concentrations are reported in pg/mL unless otherwise specified.

**Table 3:** Association of plasma biomarkers with kidney event outcomes among individuals with *APOL1* high-risk variant.

| **Biomarkers** | **N** | **N (%) with outcome** | **Model 1 HR (95% CI)** | **Model 2 HR (95% CI)** | **Model 3 HR (95% CI)** |
| --- | --- | --- | --- | --- | --- |
| **suPAR in pg/mL** |  |  |  |  |  |
| Continuous (per doubling) | 479 | 75 | 2.2 (1.5 - 3.1) | 1.9 (1.3 – 3.0) | 1.7 (1.0 – 2.6) |
| Tertiles |  |  |  |  |  |
| ≤ 2032 | 159 | 17 (22.7) | REF | REF | REF |
| > 2032 to < 2600 | 160 | 22 (29.3) | 1.3 (0.7 - 2.5) | 1.2 (0.6 – 2.3) | 1.2 (0.6 – 2.2) |
| ≥ 2600 | 160 | 36 (48.0) | 2.4 (1.3 - 4.3) | 1.8 (1.0 – 3.4) | 1.5 (0.8 – 2.8) |
| **IL-18 in pg/mL** |  |  |  |  |  |
| Continuous (per doubling) | 498 | 80 | 1.5 (1.2 - 1.9) | 1.5 (1.2 – 1.8) | 1.4 (1.1 – 1.8) |
| Tertiles |  |  |  |  |  |
| ≤ 264 | 165 | 21 (26.3) | REF | REF | REF |
| > 264 to < 415.5 | 167 | 20 (25.0) | 1.0 (0.5 - 1.8) | 1.0 (0.5 – 1.8) | 1.1 (0.6 – 2.1) |
| ≥ 415.5 | 166 | 39 (48.8) | 2.2 (1.3 - 3.7) | 2.1 (1.3 – 3.7) | 1.8 (1.0 – 3.2) |
| **YKL-40 in pg/mL** |  |  |  |  |  |
| Continuous (per doubling) | 498 | 80 | 1.4 (1.2 - 1.5) | 1.2 (1.1 – 1.4) | 1.1 (1.0 – 1.3) |
| Tertiles |  |  |  |  |  |
| ≤ 23800 | 166 | 14 (17.5) | REF | REF | REF |
| > 23800 to < 50400 | 166 | 23 (28.8) | 1.6 (0.8 - 3.2) | 1.3 (0.7 – 2.7) | 1.3 (0.6 – 2.6) |
| ≥ 50400 | 166 | 43 (53.8) | 3.5 (1.9 - 6.3) | 2.1 (1.1 – 4.1) | 1.6 (0.8 – 3.1) |
| **MCP-1 in pg/mL** |  |  |  |  |  |
| Continuous (per doubling) | 498 | 80 | 1.4 (1.2 - 1.7) | 1.4 (1.1 – 1.7) | 1.3 (1.0 – 1.6) |
| Tertiles |  |  |  |  |  |
| ≤ 124.5 | 166 | 18 (22.5) | REF | REF | REF |
| > 124.5 to < 177.5 | 166 | 21 (26.3) | 1.2 (0.6 - 2.2) | 0.9 (0.5 – 1.7) | 0.9 (0.5 – 1.7) |
| ≥ 177.5 | 166 | 41 (51.3) | 2.3 (1.3 - 4.1) | 1.6 (0.9 – 2.9) | 1.2 (0.6 – 2.1) |
| **TNFR-1 in pg/mL** |  |  |  |  |  |
| Continuous (per doubling) | 498 | 80 | 2.6 (1.8 – 3.6) | 2.2 (1.5 – 3.3) | 2.1 (1.3 – 3.2) |
| Tertiles |  |  |  |  |  |
| ≤ 2139 | 166 | 11 (6.6) | REF | REF | REF |
| 2140 to 2936 | 166 | 24 (14.5) | 2.0 (1.0 – 4.2) | 1.8 (0.9 – 3.7) | 1.7 (0.8 – 3.5) |
| ≥ 2937 | 166 | 45 (27.1) | 4.2 (2.1 – 8.0) | 3.3 (1.6 – 6.8) | 2.4 (1.1 – 4.9) |
| **TNFR-2 in pg/mL** |  |  |  |  |  |
| Continuous (per doubling) | 498 | 80 | 1.6 (1.4 – 1.9) | 1.6 (1.3 – 2.0) | 1.7 (1.3 – 2.1) |
| Tertiles |  |  |  |  |  |
| ≤ 3532 | 166 | 15 (9) | REF | REF | REF |
| 2533 to 5018 | 166 | 23 (13.9) | 1.6 (0.8 – 3.1) | 1.4 (0.7 – 2.7) | 1.4 (0.7 – 2.7) |
| ≥ 5019 | 166 | 42 (25.3) | 3.1 (1.7 – 5.6) | 2.4 (1.3 – 4.7) | 1.9 (1.0 – 3.7) |
| **KIM-1 in pg/mL** |  |  |  |  |  |
| Continuous (per doubling) | 498 | 80 | 1.8 (1.5 – 2.1) | 1.7 (1.4 – 2.1) | 1.4 (1.1 – 1.8) |
| Tertiles |  |  |  |  |  |
| ≤ 111 | 166 | 11 (6.6) | REF | REF | REF |
| 112 to 215 | 166 | 18 (10.8) | 1.8 (0.8 – 3.8) | 1.4 (0.6 – 3.1) | 1.1 (0.5 – 2.4) |
| ≥ 216 | 166 | 51 (30.7) | 6.3 (3.3 – 12.1) | 4.1 (2.0 – 8.7) | 2.3 (1.1 – 5.1) |

Model 1: Unadjusted
Model 2: Adjusted for age, sex, baseline eGFR, BMI
Model 3: Model 2 + Adjusted for DM2, hypertension, coronary disease, heart failure, SBP, ACEI/ARB use, and UACR

### 3.2 Association between plasma biomarkers and kidney outcome

presents the multivariable-adjusted associations between plasma biomarkers and the composite kidney outcome. In adjusted models, each two-fold higher biomarker concentration was independently associated with an increased risk of kidney events: hazard ratio (HR) 1.7 (95% CI 1.0–2.6) for suPAR, 2.1 (95% CI 1.3–3.2) for TNFR1, 1.7 (95% CI 1.3–2.1) for TNFR2, 1.4 (95% CI 1.1–1.8) for KIM-1, and 1.4 (95% CI 1.1–1.8) for IL-18. In contrast, YKL-40 and MCP-1 demonstrated only modest associations with the outcome after adjustment for confounders, with HRs of 1.1 (95% CI 1.0– 1.3) and 1.3 (95% CI 1.0–1.6), respectively.

### 3.3 Discrimination

The bootstrapped area under the curve (AUC) for the clinical model alone in predicting kidney events was 0.79 (SE 0.02). The addition of individual biomarkers to the clinical model resulted in minimal change in AUC. However, when all biomarkers were incorporated simultaneously, the bootstrapped AUC increased to 0.81 (SE 0.02), reference in **Table 4**.

**Table 4.** Harrell’s concordance statistic table with bootstrapped AUCs.

| **Harrell's Concordance Statistic** |  |  |  |  |
| --- | --- | --- | --- | --- |
| **Source** | **Estimate** | **Standard Error** | **Concordance** | **Discordance** |
| **Clinical** | 0.7899 | 0.0232 | 20176 | 5368 |
| **Clinical + suPAR** | 0.7969 | 0.0228 | 20357 | 5187 |
| **Clinical + IL18** | 0.8021 | 0.0216 | 20489 | 5055 |
| **Clinical + YKL40** | 0.7933 | 0.0233 | 20263 | 5281 |
| **Clinical + MCP1** | 0.7945 | 0.0231 | 20295 | 5249 |
| **Clinical + TNFR1** | 0.8004 | 0.0238 | 20445 | 5099 |
| **Clinical + TNFR2** | 0.803 | 0.0231 | 20511 | 5033 |
| **Clinical + KIM1** | 0.8063 | 0.0226 | 20596 | 4948 |
| **Clinical + All biomarkers** | 0.8128 | 0.0225 | 20763 | 4781 |

**Footnote:** Harrell's concordance statistic (C-statistic) was used to assess discriminative performance of time-to-event models. The estimate reflects the probability that, for a randomly selected pair of participants, the model correctly assigns a higher predicted risk to the individual who experiences the kidney event earlier. Standard errors were obtained through bootstrap resampling. Concordant and discordant pairs represent the number of participant pairs in which predicted and observed event ordering were in agreement or disagreement, respectively.

### 3.4 Biomarker risk scores and incidence of kidney events

**Figure 2** demonstrates a clear, graded relationship between the biology-weighted biomarker risk score and the incidence of the composite kidney endpoint. Participants were distributed across five score categories, with the solid gray bars indicating the absolute number of individuals in each category and the red line depicting the proportion experiencing the composite outcome within each category. When the five categories were consolidated into three clinically interpretable strata, the model revealed marked prognostic separation. Participants classified as low risk (score categories 1–2) had a low observed event rate (7%), whereas those in the moderate-risk strata (categories 3–4) experienced a higher event rate (16%). The high-risk stratum (category 5) demonstrated the greatest enrichment for kidney events, with more than a threefold higher event proportion relative to the low-risk group (36%). This monotonic rise across strata suggests that the composite score is not driven by a single marker but rather captures the cumulative burden of abnormal biomarker signals across multiple biological domains.

**Figure 2.**
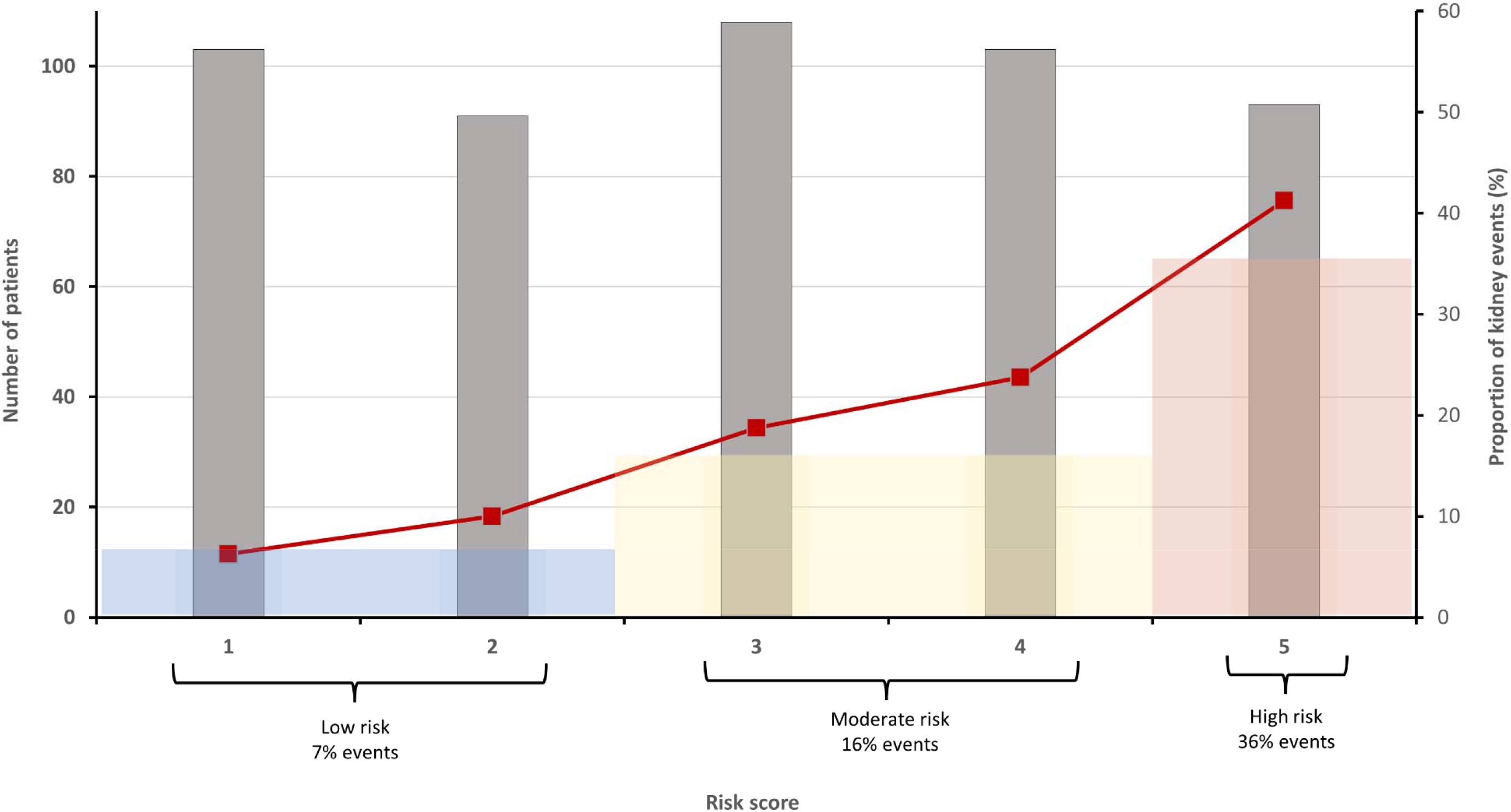
Stepwise increase in composite kidney events across biology-weighted biomarker risk score strata in individuals with high-risk APOL1 genotypes. **Footnote:** Participants were grouped into five ordered categories according to their composite biology-weighted biomarker risk score. Bars represent the number of participants (N) per category, and the line represents the proportion experiencing the composite kidney endpoint. Categories were further grouped into low-(1–2), moderate-(3–4), and high-risk (5) strata, with progressively higher event rates (7%, 16%, and 36%), demonstrating a graded relationship between cumulative biomarker burden and kidney events.

## 4. Discussion

Although high-risk *APOL1* genotypes are strongly associated with kidney disease, incomplete and variable penetrance is observed, with substantial heterogeneity in clinical expression among carriers (1–3). This paradox underscores a central limitation of genotype-based risk attribution: *APOL1* status confers susceptibility but does not define clinical trajectory. In this genetically susceptible, admixed population, plasma biomarkers reflecting inflammatory signaling and tubular injury revealed substantial heterogeneity in subsequent kidney events. When integrated into a composite, biomarker-weighted risk score, these multidomain signals identified marked differences in prognosis that were not apparent from genotype or traditional clinical measures alone.

These findings underscore an important distinction between genetic susceptibility and clinical trajectory. While *APOL1* genotype confers increased risk for kidney disease, it does not identify which individuals will experience progressive loss of kidney function (1–3). The present results suggest that circulating biomarkers may capture indirect aspects of ongoing biological processes associated with kidney injury that might not be reflected by genotype, eGFR, or albuminuria alone (12–16,20,21).

The differential strength of association observed across biomarkers is also informative. TNFR1, TNFR2, suPAR, KIM-1, and IL-18 demonstrated the strongest and most consistent associations with kidney outcomes after multivariable adjustment, supporting prior evidence that inflammatory signaling and tubular injury are central components of progressive kidney disease (12–16,20,21). In contrast, MCP-1 and YKL-40 showed more modest associations, suggesting variability in the contribution of pathways related to vascular inflammation and tissue repair within this cohort (17–19). These findings align with the emerging recognition of biological heterogeneity in *APOL1*-associated kidney disease, in which distinct pathogenic domains may predominate across individuals. For example, endothelial injury may drive one clinical phenotype, whereas primary podocyte injury—often characterized by higher levels of albuminuria—may define another. Collectively, this framework supports the concept that *APOL1*-associated kidney disease reflects the interplay of multiple biological pathways rather than a single dominant mechanism (2,5,6), and raises the possibility that future biomarker-driven approaches could enable subtype classification, refined risk prediction, and potentially more tailored therapeutic strategies.

The composite biomarker risk score further illustrated this heterogeneity. Participants categorized into low-, moderate-, and high-risk strata experienced markedly different event rates (7%, 16%, and 36%, respectively). These differences were observed in a cohort selected solely based on *APOL1* genotype and were not readily explained by traditional clinical measures. The graded, stepwise increase in event rates across risk strata supports the internal consistency of the scoring approach and suggests that the cumulative burden of abnormal biomarker signals may provide additional prognostic information (12–16,22). The consistency of associations across biomarkers and the graded risk separation observed with the composite score support the robustness of the overall framework despite multiple test comparisons.

Importantly, this work should not be interpreted as a traditional biomarker association study. Rather, it illustrates a framework for interpreting *APOL1*-associated risk through the lens of measurable disease activity. This conceptual approach parallels broader clinical precedent in which integrated biomarker-enhanced models have been used to improve risk prediction in other medical settings (10,11).

Several limitations should be acknowledged. UACR was imputed for a small subset of participants using a conservative approach and was included only as an adjustment covariate. Biomarkers were measured at a single time point, precluding assessment of temporal changes in biological activity. Observational design limits causal inference, and residual confounding is possible. The risk score was derived and tested within the same Bio*Me* cohort without external validation, which may limit generalizability. In addition, only individuals with biallelic high-risk *APOL1* variants were included, while recent evidence from Africa suggests that monoallelic carriers may also experience adverse outcomes. Although the score clearly stratified event rates, improvements in Harrell’s C-statistic were modest and may be better defined in larger, more comprehensively characterized populations.

In conclusion, among individuals with high-risk *APOL1* genotypes, plasma biomarkers of inflammation and tubular injury were independently associated with kidney outcomes and, when combined into a weighted score, identified groups with substantially different risks of progression. These findings suggest that multidomain biomarker assessment may complement traditional clinical measures in the prognostic evaluation of *APOL1*-associated kidney disease and warrant further study in external cohorts and prospective settings. Future work should evaluate model calibration in external cohorts (e.g., predicted vs observed risk at prespecified time horizons) to support clinical implementation.

## Data Availability

All data produced in the present study are available upon reasonable request to the authors

## Authors’ contributions

GVR and SGC drafted the manuscript. GVR and KC contributed equally to the development of this manuscript and share co–first authorship. KC, GVR, NN, PP, LC, GNN, and SGC provided substantial intellectual contributions to the study design, data interpretation, and manuscript revision. All authors reviewed and approved the final version of the manuscript.

## Acknowledgment

The authors gratefully acknowledge all participants enrolled in the Mount Sinai BioMe Biobank.

